# Vaccine effectiveness against SARS-CoV-2 Delta and Omicron infection and infectiousness within households in the Netherlands between July 2021 and August 2022

**DOI:** 10.1101/2023.01.10.23284386

**Authors:** Christina E. Hoeve, Brechje de Gier, Anne J. Huiberts, Hester E. de Melker, Susan J.M. Hahné, Susan van den Hof, Mirjam J. Knol

**Affiliations:** Centre for Infectious Disease Control, National Institute for Public Health and the Environment, Bilthoven, the Netherlands

## Abstract

**Introduction:** We aimed to estimate vaccine effectiveness against infection (VE- infection) and infectiousness (VE-infectiousness) in a household setting during Delta and Omicron. Knowing these effects can aid policy makers in deciding which groups to prioritize for vaccination.

**Methods:** Participants with a positive SARS-CoV-2 test were asked about COVID-19 vaccination status and SARS-CoV-2 testing of their household members one month later. VE-infection and VE-infectiousness was estimated using GEE logistic regression adjusting for age and vaccination status, calendar week and household size.

**Results:** 3,409 questionnaires concerning 4,123 household members were included. During the Delta-period, VE-infection of primary series was 47% (95% CI: −27%-78%) and VE-infectiousness of primary series was 70% (95% CI: 28%-87%). During the Omicron-period, VE-infection was −36% (95% CI: −88%-1%) for primary series and −30% (95% CI: −80%-6%) for booster vaccination. The VE-infectiousness was 45% (95% CI: −14%-74%) for primary series and 64% (95% CI: 31%-82%) for booster vaccination.

**Discussion:** Our study shows that COVID-19 vaccination is effective against infection with SARS-CoV-2 Delta and against infectiousness of SARS-CoV-2 Delta and Omicron. Estimation of VE against infection with SARS-CoV-2 Omicron was limited by several factors. Our results support vaccination for those in close contact with vulnerable people to prevent transmission.

## INTRODUCTION

COVID-19 vaccines may contribute to controlling SARS-CoV-2 infections by reducing the susceptibility of an individual of becoming infected and/or by reducing the infectiousness when a vaccinated individual is infected. The reduction in susceptibility afforded by vaccines is referred to as the vaccine effectiveness against infection (VE-infection), and the reduction in infectiousness of an individual once infected as vaccine effectiveness against infectiousness (VE-infectiousness) (1). In many studies it is difficult to determine the VE-infectiousness. Household studies have proven to be excellent tools to study VE against both infection and infectiousness. For policy makers both VE-infection and VE-infectiousness are relevant. If vaccines only prevent infections through reducing susceptibility then vaccination of only vulnerable populations such as the elderly or medical risk groups may suffice. However, if vaccines also prevent onward transmission (when infected) by reducing infectiousness then population-wide vaccination or extending the target group for vaccination, for example to health care workers, can have added value to protect those who remain at high risk of severe disease.

The effectiveness of COVID-19 vaccines against infection has been investigated extensively. It has been shown that vaccination substantially reduces the risk of SARS-CoV-2 infections, although the effect differs by variant of concern and protection decreases with increasing time since vaccination(2, 3). After its emergence in late 2020, the SARS-CoV-2 Delta variant showed increased transmissibility compared to previous variants and quickly became the dominant variant worldwide. The more recent Omicron subvariants are even more transmissible. Recent data have shown that COVID-19 vaccines can reduce the onward transmission of Omicron by infected individuals (4). It has also been reported that the infectiousness of Omicron is higher than Delta, regardless of vaccination status (5, 6).

In this study we assess the VE-infection and VE-infectiousness of the SARS-CoV-2 Delta and Omicron variants for vaccines authorized in the Netherlands during the second half of 2021 and the first half of 2022.

## METHODS

### Study design

VASCO (Vaccine Study COVID-19) is a prospective cohort study of ∼45,000 community-dwelling Dutch adults between 18-85 years, which aims to estimate long-term effectiveness of COVID-19 vaccines that have been used in the Dutch national COVID-19 vaccination program (7). Enrollment of participants took place between 3 May 2021 and 15 December 2021 and participants are followed for five years. The VASCO study population is relatively old with a median age of 61 years due to oversampling of older adults. Participants are followed with monthly questionnaires for COVID-19 vaccination and self-reported positive SARS-CoV-2 tests, and fingerprick samples were requested at baseline, 6 months and 12 months follow-up. Participants can report a positive SARS-CoV-2 test (PCR or (self-administered) antigen test) directly in a mobile phone application designed for the study, including date and type of test. The study provided antigen self-tests to participants free-of-charge from April 2022 onwards because from that date onwards, national test centers no longer offered testing free-of-charge to the general public. From the end of August 2021, participants who reported a positive SARS-CoV-2 test were asked one month after the positive test date to complete an additional questionnaire about their household members. Household member data included age, whether or not they were tested for SARS-CoV-2 in the window period (two weeks before to four weeks after the positive test of the VASCO participant), the date of the test, the result of the test (positive or negative), and the number, dates and types of COVID-19 vaccinations received. The participant was asked whether the household member gave verbal permission for reporting their infection and vaccination status.

### Study period

The study period started on 23 July 2021, which was the date of the first infection of one of the participants for which a questionnaire on household members was completed, and ended on 7 August 2022 when the last infection in the household sub-study was reported. The study period therefore included the time periods in which the Delta variant and the Omicron BA.1, BA.2, BA.4 and BA.5 subvariants were dominant in the Netherlands (8). A variant was considered dominant if more than 90% of the SARS-CoV-2 samples that were sequenced as part of the national pathogen surveillance program detected that variant. The Delta-dominant period was from the start of the study period until 18 December 2021. The Omicron-dominant period was from 11 January 2022 until the end of the study period. The BA1, BA2 and BA4/BA5 subvariant dominant periods were from 9 January 2022 – 31 January 2022, 20 March 2022 – 16 May 2022 and 27 June 2022 – end of study period, respectively.

### Inclusion and exclusion criteria

Only households in which the VASCO participant was classified as the index case were included in the current analysis, defined as the participant being the first in the household to test positive according to the questionnaire. Households were excluded if the index case had a previous positive SARS-CoV-2 test before the index case date or if antibodies against the SARS-CoV-2 nucleocapsid protein (anti-N) were present in any blood sample before the index case date. The index case date was defined as the reported date of the positive SARS-CoV-2 test (PCR or antigen (self-)test). Households with co-primary cases were excluded from the analysis since in these situations an index could not be clearly defined. Co-primary cases were defined as index and household members who tested positive within one day. We could not use onset date of symptoms to define co-primary cases as this date was not available for household members. Households in which at least one member did not give permission for sharing their data were excluded from the analysis. Household members who did not do a SARS-CoV-2 test in the window period were excluded from the analysis. Information on previous infections from household members was unavailable. A secondary case is defined as a household member who is reported to have had a positive test for SARS-COV-2 within 2-14 days after the index case date.

### Vaccination status

Vaccination status was determined for both index cases as well as household members on the index case date. Information on vaccination was self-reported through the questionnaires. For index cases additional information on vaccinations until March 2022 was available from the national vaccination register (CIMS). In the Netherlands, informed consent of the vaccinee is required for a COVID-19 vaccination to be registered in CIMS; this registry is therefore incomplete. Vaccination data from the registry was used, except when no informed consent was given for registration of any of the received vaccines in CIMS or no consent was given for linking study data with CIMS data. If only part of the vaccination data was available in CIMS and/or self-reported, data was combined (9). A person was considered unvaccinated if no vaccine was registered or reported. A person was considered vaccinated with a primary series if a second dose of Comirnaty (BNT162b2; BioNTech/Pfizer, Mainz, Germany/New York, United States (US)), Spikevax (mRNA-1273, Moderna, Cambridge, US), Vaxzevria (ChAdOx1-S; AstraZeneca, Cambridge, United Kingdom) was received at least 14 days before the index case date or if a first dose of Jcovden (Ad26.COV2-S (recombinant), Janssen-Cilag International NV, Beerse, Belgium) was received at least 28 days before the index case date. A person was considered to be vaccinated with a first booster if a third dose of Comirnaty, Spikevax or Vaxzevria was received at least 7 days before the index case date. A third dose administered before the start of the booster campaign (18 November 2021) was considered an additional primary series vaccination, and was only offered to persons with a severe immune deficiency, and were thus not considered booster doses. If a person was vaccinated with Jcovden and received a second dose more than 90 days after the first dose, then this person was considered to be vaccinated with a first booster if the second dose was received at least 7 days before the index case date. The vaccination status for the second booster was the same as for the first booster with an additional vaccination. Index cases or household members who had a vaccination status other than unvaccinated, primary series, or boosted were excluded.

### Serology

Fingerprick samples were analyzed with the Elecsys anti-N assays on the Cobas e801 (Roche Diagnostics, Mannheim, Germany), which are electrochemiluminescence immunoassays measuring Ig levels against the SARS-CoV-2 nucleocapsid protein (anti-N antibodies). The qualitative cut-off index (COI) was converted to numeric results in BAU/ml using batch-specific, linear calibration-lines obtained with a dilution range of the NIBSC 20/136 WHO standard (NIBSC). The cut-off for anti-N-positivity was set by converting COI 1.0 to corresponding BAU/ml using these calibration lines.

### Data analysis

Characteristics of index cases and household members are presented with frequencies and percentages. The time to secondary infection is calculated as the difference between positive test date of the index and the household member. Time to secondary infection is reported as median and interquartile range. Secondary attack rates (SAR) are calculated as the proportion of household members who test positive for SARS-CoV-2 after the index case date. SAR are stratified by vaccination status and SARS-CoV-2 variant and compared using Chi-squared tests. VE-infection during the Delta- and Omicron-dominant periods was estimated using logistic regression with and without adjustment for the age group of the household member (0-17, 18-39, 40-59, 60+), the age group of the index case (18-39, 40-59, 60-85), calendar week as a categorical variable, vaccination status of the index and household size. VE-infectiousness during the Delta- and Omicron-periods was estimated using logistic regression with and without adjustment for age group of the index case (18-39, 40-59, 60-85), age group of the household member (0-17, 18-39, 40-59, 60+), vaccination status of the household member, calendar week as categorical variable, and household size. Generalized estimating equations models with exchangeable correlation structure were used to take into account dependencies within the household. In a sensitivity analysis we stratified vaccination status based on time since vaccination (<90 days and >= 90 days). In a further sensitivity analysis, households with index cases with a prior infection were included in the analysis to evaluate the impact of prior infection on the VE-infectiousness. A third sensitivity analysis explored the impact of age on VE-infection and VE-infectiousness by excluding all household members under 18 years, since people under 18 are often less vaccinated. In a fourth sensitivity analysis we reduced the time window for testing of household members to 7 days instead of 14. The results of the sensitivity analyses are presented in the supplementary files. All data processing and statistical analyses were done in R version 4.0.2 using package geepack for the VE analyses.

### Ethical statement

The VASCO study protocol was approved by the not-for-profit independent Medical Ethics Committee of the *Stichting Beoordeling Ethiek Biomedisch Onderzoek* (BEBO), Assen, the Netherlands (NL76815.056.21) (7). Written informed consent was obtained from all participants prior to enrollment into the study (7). For linking of national registration of COVID-19 vaccination data participants could choose to consent or not (7). The participant should indicate for each household member whether or not they gave permission for sharing of their data. If no permission was indicated for a household member then this entire household was excluded from the analysis. Further details on data management, privacy and ethics regarding the VASCO study are described by Huiberts et al (7).

## RESULTS

For infections reported between 23 July 2021 and 7 August 2022, 13,138 household questionnaires were completed by VASCO participants. Of these, 3,409 questionnaires (25.9%) concerning 4,123 household members were included in the analysis (**figure 1**). The characteristics of index cases and household members are presented in **table 1**. The median age of index cases was 61 years (range 18-85). At the index case date, 2% of the index cases was unvaccinated, 10% had completed a primary vaccination series, 76% had received a first booster and 12% a second booster. A total of 4,123 household members were included with a median age of 58 years (range 0-93). At the index case date, 11% of the household members was unvaccinated, 25% had completed a primary vaccination series, 55% had received a first booster and 9% a second booster. The median time to secondary infection was 4.5 days (IQR: 3-6 days) during the Delta period and 4.0 days (IQR: 3-6 days) during the Omicron period (**figure 2**). The proportion of household members who tested fluctuated per month, with 75-100% during Delta period and 75%-88% during Omicron period.

**Table 1.** Characteristics of index cases and household members.

|  | N (column %) |  |
| --- | --- | --- |
| Characteristic | Index,<br>3,409 | Household members,<br>4,123 |
| **Age group** |  |  |
| Median (IQR) | 61 (50 - 65) | 58 (37 - 65) |
| 0-17 | - | 527 (13%) |
| 18-39 | 423 (12%) | 658 (16%) |
| 40-59 | 1119 (33%) | 1028 (25%) |
| 60+ | 1867 (55%) | 1910 (46%) |
| **Gender** |  |  |
| Male | 1,306 (38%) | Not available |
| Female | 2,101 (62%) | Not available |
| Other | 2 (0%) | Not available |
| **COVID-19 vaccination status at index case date** |  |  |
| Not vaccinated | 57 (2%) | 462 (11%) |
| Primary series | 328 (10%) | 1,043 (25%) |
| Booster1 | 2,601 (76%) | 2,250 (55%) |
| Booster2 | 423 (12%) | 368 (9%) |
| **COVID-19 case status** |  |  |
| Yes | 3,409 (100%) | 1,811 (44%) |
| No | 0 (0%) | 2,309 (56%) |
| Unknown | 0 (0%) | 3 (0%) |
| **Number of household members** |  |  |
| 1 | 2,596 (76%) | 2,596 (63%) |
| 2 | 403 (12%) | 621 (15%) |
| 3 | 318 (9%) | 659 (16%) |
| 4 | 83 (2%) | 209 (5%) |
| 5 | 8 (0%) | 32 (1%) |
| 6 | 1 (0%) | 6 (0%) |

**Figure 1.**
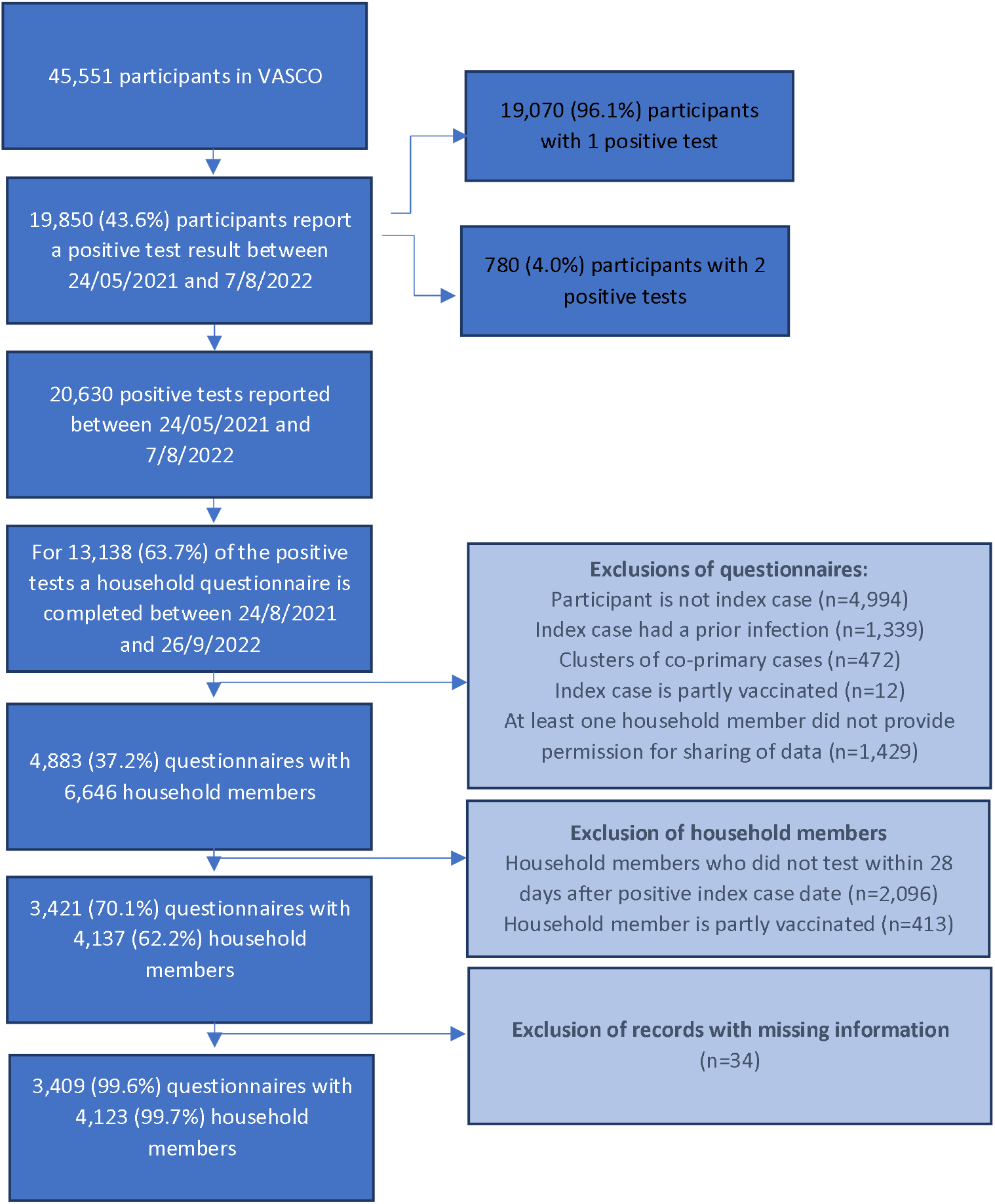
In- and exclusion of questionnaires and household members.

**Figure 2.**
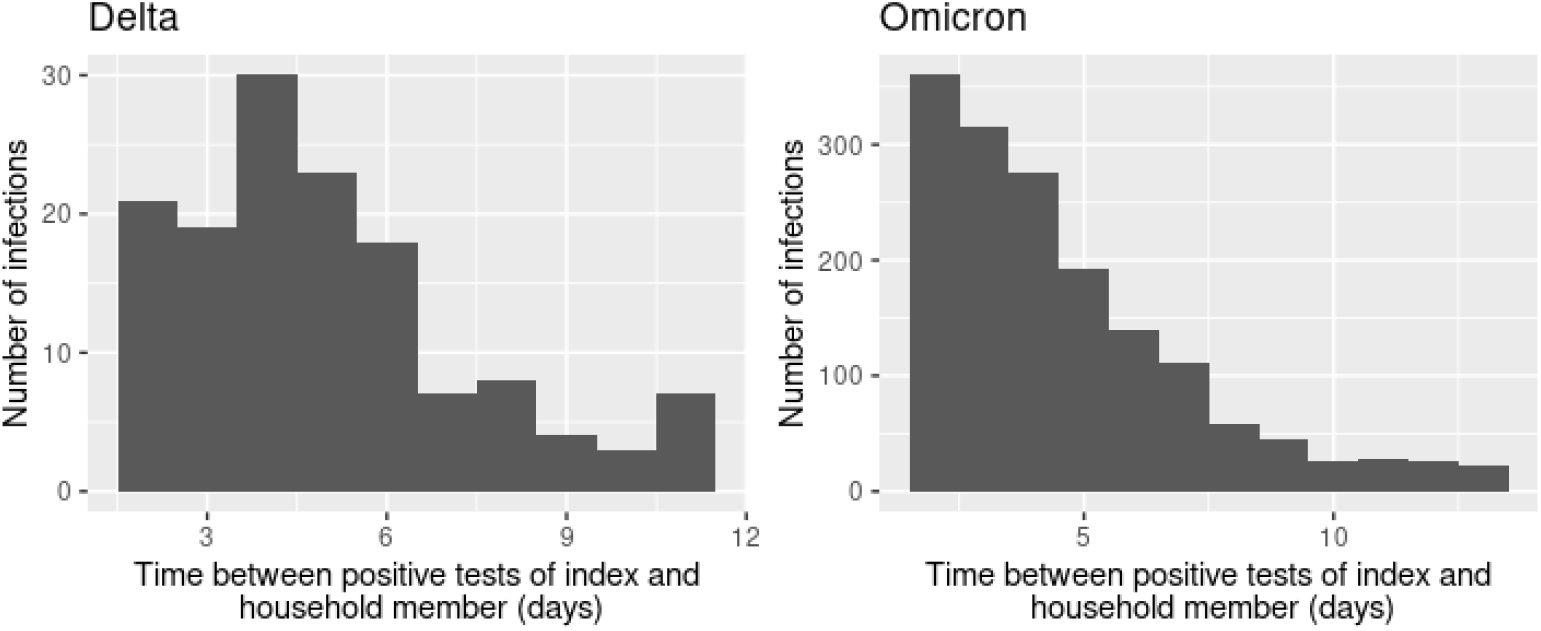
Distribution of time to secondary infection within contributing households.

The overall SAR was almost the same during the Omicron period (43%, 95% CI:41%;44%) as during the Delta period (41%, 95% CI:36%;47%). The SAR was not significantly different during the Omicron subvariant periods: 41% (95% CI: 34%;48%) during the BA.1 period, 37% during the BA.2 period (95% CI: 33%;40%) and 44% during the BA.4/5 period (95% CI: 40%;48%). The SAR in households with unvaccinated index cases was not different during Delta (55%, 95% CI:38%;72%) and Omicron periods (57%, 95% CI:43%;71%). Although numbers in subgroups are generally low, the SAR showed a decrease in households where the index case had received primary or booster vaccinations, both during Delta and Omicron periods (**supplementary data, table A**).

During the Delta-dominant period the adjusted VE-infection was 47% (95% CI:-27%;78%) for household members with a primary series. There were not enough household members with booster vaccinations to obtain reliable estimates. During the Omicron period no direct protection against infection was found as the adjusted VE-infection was −36% (95% CI:-88%;1%) for household members with a primary series and −30% (95% CI:-80%;6%) for household members with a first booster vaccination (**table 2**). VE-infection for the second booster could not be reliably estimated since the majority of unvaccinated individuals was under 60 and the majority of individuals with a second booster was over 60 years, resulting in a faulty comparison.

**Table 2.** Vaccine effectiveness against infection. Impact of the vaccination status of the household member on contracting infection.

| Variant | Vaccination status household member | Model 1: VE adjusted for calendarweek and vaccinationstatus index | Model 2: Model 1 + household size + age household member + age index | Number of household members |
| --- | --- | --- | --- | --- |
| Delta | No vaccination | Ref | Ref | 94 |
|  | Primary series | 24% (-40%-59%) | 47% (-27%-78%) | 243 |
| Omicron | No vaccination | Ref | Ref | 364 |
|  | Primary series | -41% (-85%--7%) | -36% (-88%-1%) | 771 |
|  | Booster 1 | -58% (-102%--24%) | -30% (-80%-6%) | 2233 |

During the Delta period, the adjusted VE-infectiousness was 70% (95% CI:28%;87%) for index cases with a primary series and 93% (95% CI:56%;99%) for index cases with a booster vaccination. There were only 12 index cases with a booster during the Delta period and the booster was administered at the end of the Delta period, therefore this estimate should be interpretated with caution. During the Omicron period the adjusted VE-infectiousness was 45% (95% CI:-14%;74%) for index cases with a primary series and 64% (95% CI:31%;82%) for index cases with a first booster vaccination (**table 3**).

**Table 3.**
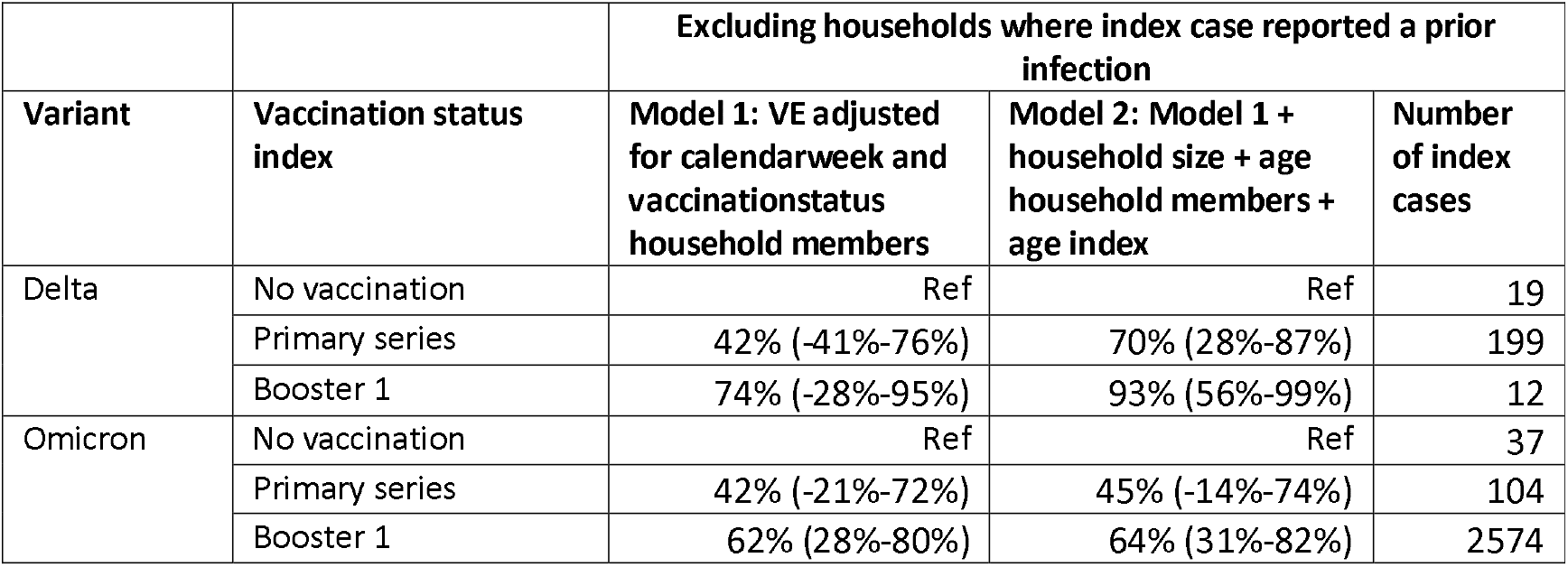
Vaccine effectiveness against infectiousness. Impact of the vaccination status of the index case on transmission to household members.

In a sensitivity analysis we explored the impact of a prior SARS-CoV-2 infection on the VE-infectiousness. This sensitivity analysis did not evaluate VE-infection as information on prior infection among household members was not available. Both during Delta and Omicron periods we observed a small decrease in the VE-infectiousness when including index cases with a prior infection. VE-infectiousness during Delta was estimated at 61% (95% CI:3%;84%) for primary series and 85% (95% CI:10%;98%) for first booster vaccination. During Omicron this was 32% (95% CI:-12%;59%) for primary series and 32% (95% CI:-5%;56%) for first booster vaccination (**supplements, table B**).

A sensitivity analysis exploring the effect of time since vaccination showed that VE estimates were lower after three months since vaccination compared to the first three months since vaccination (**supplements, table C**). A sensitivity analysis exploring the impact of younger household members showed an increase in VE-infectiousness and a decrease in VE-infection (**supplements, table D**). Another sensitivity analysis exploring the impact of the time window for testing showed similar estimates as to the main analysis (**supplement table E1 and E2)**.

## DISCUSSION

In this prospective cohort study among adults aged 18-85 years we collected data on infections in household members given infection in the household to estimate VE-infectiousness and VE-infection during the Delta and Omicron dominant periods. We found an adjusted VE-infectiousness of 70% and 93% during the Delta period for primary series and first booster respectively. VE-infectiousness was lower during the Omicron period: 45% and 64% for primary series and first booster respectively. We found a VE-infection of 47% for primary series during the Delta period. During the Omicron period the VE-infection was −36% and −30% for primary series and first booster respectively.

This study shows that COVID-19 vaccination is effective against infectiousness of SARS-CoV-2 Delta and Omicron in household settings for index cases without prior infection, both for indexes vaccinated with a primary series and indexes with a booster in line with other studies (4, 6). Another study from the Netherlands, that studied VE of primary vaccination against infectiousness during the Delta period using source and contact tracing data, showed fairly similar results with a VE-infectiousness of 63% (46%-75%) for unvaccinated contacts and 40% (20%-54%) for contacts with a primary series (10). A study by Jalali et al. showed that the risk of onward transmission of a Delta infection to household members was 82% (RR: 0.18; 95% CI: 0.01-0.70) lower for booster vaccinated index cases compared to unvaccinated index cases, although no difference was observed for Omicron (RR: 0.99; 95% CI: 0.68-1.49) (5). In contrast, another study reported no difference in VE-infectiousness between Delta and Omicron with an overall increased odds ratio of 1.41 (95% CI: 1.27-1.57) for infectiousness of unvaccinated index cases compared to index cases with a primary series (6). It should be noted that the studies by Jalali and Lyngse were done in the early Omicron waves and likely concern mostly BA.1 variants, whereas our estimates also include infections with Omicron BA.2 and BA.4/5 variants. Both studies excluded index cases with a prior infection, although this was only based on lab-confirmed tests, whereas we also excluded prior infections based on self-tests and presence of SARS-CoV-2 anti-N antibodies. In a sensitivity analysis we included index cases with a prior infection which led to a decrease in the VE-infectiousness, suggesting that prior infection gives protection against infectiousness thereby diluting the VE estimates. Earlier reports also showed that index cases with a prior infection had a lower risk of transmitting to close contacts (4).

A lower VE against Omicron infection compared to Delta has been observed in many other studies (5, 6, 11). However, the estimates we found on VE against Omicron infection are much lower than found in other studies. This could be due to several reasons. First, we could not adjust for prior infections of household members as this information was not available. Prior infections would reduce susceptibility among household members, diluting vaccine effects in our analysis. Second, the VE-infection in this study is in the context of exposure to infection in a household setting. Close contact of long duration in a household setting might have resulted in increased infection risk, even when vaccinated. Symptomatic infection has been linked to increased infectiousness compared to asymptomatic infection (12). Since the vast majority (96%) of our index cases had a symptomatic infection this might also have lowered the VE estimates. Third, time since vaccination could have played a role. In a sensitivity analysis we showed that a large proportion of participants and household members had received their last vaccination more than three months ago, which will have led to a reduction in the VE. Finally, since age was a factor influencing vaccination we investigated whether this could have impacted our VE estimates. A sensitivity analysis in which household members under 18 years were excluded resulted in a further reduction of the VE-infection. Therefore an imbalance of vaccination over age groups does not explain our low estimates of VE-infection.

One of the strengths of this study is that we collected data on a relatively large number of households, which enabled us to estimate VE-infectiousness. Also, we were able to assess VE during the later Omicron wave since participants in VASCO could apply for free-of-charge self-test kits as of April 2022 when national testing was scaled down. Our study also has some limitations. First, differences in testing behavior and moment of testing between vaccinated and unvaccinated index cases and household members may have affected our estimates in both directions. For example, vaccinated individuals may be more likely to test because they are more health conscious. On the other hand, they may be less likely to test because they feel safe or have no or less symptoms. Also, at some times during the study period, there were different quarantine rules for vaccinated and unvaccinated household members which could have increased differential testing behavior. For example, during the first months of the study period people vaccinated with a primary series did not have to quarantine if a household member was positive, but unvaccinated people did. We tried to limit the effects of testing behavior by only including household members for which it was reported that they did a test, although also the number of tests and the timing of testing could have been different. Second, we assumed that all infections that occurred in the household between 2-14 days after the index case were caused by the index case, while these infections could also have another origin. Misclassification of infection origin would have resulted in an underestimation of the VE-infectiousness. To minimize this we used a follow-up period of maximum 14 days. A sensitivity analysis using a follow-up period of maximum 7 days gave similar estimates (**supplement table E1 and E2**). Third, we did not collect data on the behavioral response of household members following the positive test result of the index case. Potentially, unvaccinated household members may have kept more distance than vaccinated household members, thereby lowering their risk of infection. If this is the case this would have led to an underestimation of the VE-infectiousness. Fourth, our data is based on self-report, except for the vaccination status of the index case which was verified by the national vaccination register, which could have resulted in recall bias. However, given the short time between infection of the index date and filling out the questionnaire on household members (median of 32 days) and the impact the event probably had on the household, recall bias is expected to be minimal.

## Conclusion

We showed that COVID-19 vaccination is effective against infection with SARS-CoV-2 Delta and against infectiousness of SARS-CoV-2 Delta and Omicron. Therefore, our results support vaccination especially for those in close contact with vulnerable people, such as healthcare workers, employees in elderly care homes or people otherwise often in close contact with vulnerable people and persons not eligible for vaccination.

## Data Availability

The data produced in this manuscript are available in aggregated and anonymized form upon reasonable request to the authors.

