## Supplements for "Vaccine effectiveness against SARS-CoV-2 Delta and Omicron infection and infectiousness within households in the Netherlands between July 2021 and August 2022"

**Supplement Table A. Secondary attack rates (SAR) by vaccination status of index cases and household members overall and stratified by SARS-CoV-2 variant.**

|  | **Overall** | | **Delta** | | **Omicron** | |
| --- | --- | --- | --- | --- | --- | --- |
|  | **No./total** | **SAR % (95% CI)** | **No./total** | **SAR % (95% CI)** | **No./total** | **SAR % (95% CI)** |
| **Overall** | 1761/4123 | 43 (41-44) | 140/338 | 41 (36-47) | 1601/3736 | 43 (41-44) |
| **Vaccination status index** |  |  |  |  |  |  |
| Not vaccinated | 46/83 | 55 (45-66) | 18/33 | 55 (38-72) | 28/49 | 57 (43-71) |
| Primary series | 202/465 | 43 (39-48) | 117/291 | 40 (35-46) | 70/142 | 49 (41-58) |
| Booster 1 | 1304/3111 | 42 (40-44) | 3/12 | 25 (1-50) | 1296/3084 | 42 (40-44) |
| **Vaccination status household members** |  |  |  |  |  |  |
| Not vaccinated | 187/462 | 40 (36-45) | 47/94 | 50 (40-60) | 138/364 | 38 (33-43) |
| Primary series | 412/1043 | 40 (37-42) | 93/243 | 38 (32-44) | 305/771 | 40 (36-43) |
| Booster 1 | 974/2250 | 43 (41-45) | 0/1 | 0 (0-0) | 970/2233 | 43 (41-45) |

|  | **BA1** | | **BA2** | | **BA4/BA5** | |
| --- | --- | --- | --- | --- | --- | --- |
|  | **No./total** | **SAR % (95% CI)** | **No./total** | **SAR % (95% CI)** | **No./total** | **SAR % (95% CI)** |
| **Overall** | 76/184 | 41 (34-48) | 312/852 | 37 (33-40) | 252/572 | 44 (40-48) |
| **Vaccination status Index** |  |  |  |  |  |  |
| Not vaccinated | 5/10 | 50 (19-81) | 3/7 | 43 (6-80) | 0/3 | 0 (0-0) |
| Primary series | 12/27 | 44 (26-63) | 11/19 | 58 (36-80) | 6/13 | 46 (19-73) |
| Booster 1 | 58/140 | 41 (33-50) | 266/744 | 36 (32-39) | 164/383 | 43 (38-48) |
| **Vaccination status household members** |  |  |  |  |  |  |
| Not vaccinated | 15/28 | 54 (35-72) | 13/49 | 27 (14-39) | 14/49 | 29 (16-41) |
| Primary series | 22/56 | 39 (26-52) | 57/174 | 33 (26-40) | 37/113 | 33 (24-41) |
| Booster 1 | 39/100 | 39 (29-49) | 212/548 | 39 (35-43) | 103/239 | 43 (37-49) |

**Supplement Table B. Vaccine effectiveness against infectiousness after including index cases with a prior infection.** Impact of the vaccination status of the index case on transmission to household members.

| **Variant** | **Vaccination status index** | **Model 1: VE adjusted for calendarweek and vaccinationstatus household members** | **Model 2: Model 1 + household size + age household members + age index** | **Number of index cases** |
| --- | --- | --- | --- | --- |
| Delta | No vaccination |  | Ref | 21 |
|  | Primary series | 32% (-66%-72%) | 61% (3%-84%) | 205 |
|  | Booster 1 | 59% (-83%-91%) | 85% (10%-98%) | 13 |
| Omicron | No vaccination |  | Ref | 83 |
|  | Primary series | 35% (-7%-60%) | 32% (-12%-59%) | 204 |
|  | Booster 1 | 35% (0%-58%) | 32% (-5%-56%) | 2953 |

**Supplement Table C1. Vaccine effectiveness against infection stratified by time since vaccination.** Impact of the vaccination status of the household member on contracting infection.

|  |  | **<3 months since vaccination** | | **>3 months since vaccination** | |
| --- | --- | --- | --- | --- | --- |
| **Variant** | **Vaccination status household member** | **VE adjusted for calendarweek and vaccinationstatus household member + household size + age household member + age index** | **Number of household members** | **VE adjusted for calendarweek and vaccinationstatus household member + household size + age household member + age index** | **Number of household members** |
| Delta | No vaccination | Ref | 94 | Ref | 94 |
|  | Primary series | 92% (66%-98%) | 72 | 12% (-138%-67%) | 157 |
| Omicron | No vaccination | Ref | 364 | Ref | 364 |
|  | Primary series | -30% (-103%-17%) | 205 | -24% (-78%-14%) | 566 |
|  | Booster 1 | -3% (-51%-29%) | 1549 | -62% (-145%--8%) | 684 |

**Supplement Table C2. Vaccine effectiveness against infectiousness stratified by time since vaccination.** Impact of the vaccination status of the index case on transmission to household members.

|  |  | **<3 months since vaccination** | | **>3 months since vaccination** | |
| --- | --- | --- | --- | --- | --- |
| **Variant** | **Vaccination status household member** | **VE adjusted for calendarweek and vaccinationstatus household member + household size + age household member + age index** | **Number of index cases** | **VE adjusted for calendarweek and vaccinationstatus household member + household size + age household member + age index** | **Number of index cases** |
| Delta | No vaccination | Ref | 16 | Ref | 17 |
|  | Primary series | 83% (39%-95%) | 81 | 68% (10%-89%) | 164 |
| Omicron | No vaccination | Ref | 33 | Ref | 33 |
|  | Primary series | 64% (19%-84%) | 65 | 32% (-49%-69%) | 74 |
|  | Booster 1 | 66% (32%-83%) | 1740 | 58% (15%-79%) | 1128 |

**Supplement Table D1. Vaccine effectiveness against infection including only household members older than 18 years.** Impact of the vaccination status of the household member on contracting infection.

| **Variant** | **Vaccination status household member** | **Model 1: VE adjusted for calendarweek and vaccinationstatus index** | **Model 2: Model 1 + household size + age household member + age index** | **Number of house-hold member** |
| --- | --- | --- | --- | --- |
| Delta | No vaccination |  | Ref | 26 |
|  | Primary series | 21% (-101%-69%) | 30% (-120%-80%) | 224 |
| Omicron | No vaccination |  | Ref | 104 |
|  | Primary series | -44% (-135%-11%) | -47% (-140%-10%) | 610 |
|  | Booster 1 | -53% (-145%-4%) | -42% (-128%-12%) | 2218 |

**Supplement Table D2. Vaccine effectiveness against infectiousness including only household members older than 18 years.** Impact of the vaccination status of the index case on transmission to household members.

| **Variant** | **Vaccination status index** | **Model 1: VE adjusted for calendarweek and vaccinationstatus household members** | **Model 2: Model 1 + household size + age household members + age index** | **Number of index cases** |
| --- | --- | --- | --- | --- |
| Delta | No vaccination |  | Ref | 17 |
|  | Primary series | 65% (-34%-91%) | 90% (40%-100%) | 193 |
|  | Booster 1 | 84% (-1%-98%) | 100% (70%-100%) | 12 |
| Omicron | No vaccination |  | Ref | 33 |
|  | Primary series | 51% (-19%-80%) | 57% (-6%-82%) | 100 |
|  | Booster 1 | 73% (38%-78%) | 76% (45%-89%) | 2490 |

**Supplement Table E1. Vaccine effectiveness against infection including only household members tested within 7 days of the index case date.** Impact of the vaccination status of the household member on contracting infection.

| **Variant** | **Vaccination status household member** | **Model 1: VE adjusted for calendarweek and vaccinationstatus index** | **Model 2: Model 1 + household size + age household member + age index** | **Number of house-hold member** |
| --- | --- | --- | --- | --- |
| Delta | No vaccination |  | Ref | 94 |
|  | Primary series | -27% (-154%-37%) | 25% (-104%-73%) | 243 |
| Omicron | No vaccination |  | Ref | 364 |
|  | Primary series | -45% (-93%--9%) | -25% (-77%-12%) | 771 |
|  | Booster 1 | -72% (-124%--32%) | -20% (-72%-16%) | 2233 |

**Supplement Table E2. Vaccine effectiveness against infectiousness including only household members tested within 7 days of the index case date.** Impact of the vaccination status of the index case on transmission to household members.

| **Variant** | **Vaccination status index** | **Model 1: VE adjusted for calendarweek and vaccinationstatus household members** | **Model 2: Model 1 + household size + age household members + age index** | **Number of index cases** |
| --- | --- | --- | --- | --- |
| Delta | No vaccination |  | Ref | 19 |
|  | Primary series | 51% (-38%-83%) | 72% (24%-90%) | 199 |
|  | Booster 1 | 71% (-56%-95%) | 91% (45%-99%) | 12 |
| Omicron | No vaccination |  | Ref | 37 |
|  | Primary series | 16% (-75%-60%) | 13% (-84%-59%) | 104 |
|  | Booster 1 | 54% (10%-76%) | 51% (4%-75%) | 2574 |
